# NeuroSmog: Determining the impact of air pollution on the developing brain: project protocol

**DOI:** 10.1101/2021.10.22.21265366

**Authors:** Iana Markevych, Natasza Orlov, James Grellier, Katarzyna Kaczmarek-Majer, Małgorzata Lipowska, Katarzyna Sitnik-Warchulska, Yarema Mysak, Clemens Baumbach, Maja Wierzba-Łukaszyk, Munawar Hussain Soomro, Mikołaj Compa, Bernadetta Izydorczyk, Krzysztof Skotak, Anna Degórska, Jakub Bratkowski, Bartosz Kossowski, Aleksandra Domagalik, Marcin Szwed

**Author notes:** **Corresponding author** Marcin Szwed, PhD, Institute of Psychology, Jagiellonian University, Ingardena 6, 30-060 Krakow, Poland.

## Abstract

**Background:** Exposure to airborne particulate matter (PM) may affect neurodevelopmental outcomes in children. The mechanisms underlying these relationships are not currently known. We aim to assess whether PM affects the developing brains of schoolchildren in Poland, a European country characterized by very high levels of particulate air pollution.

**Methods:** Between 2020 and 2022, 800 children aged 10 to 13 years are being recruited as participants in a case-control study. Cases (children with attention deficit hyperactivity disorder (ADHD)) are being recruited from psychology clinics. Population-based controls are being sampled from schools. The study area comprises 18 towns in southern Poland characterized by wide-ranging levels of PM. Comprehensive psychological assessments are being conducted to assess cognitive and social functioning. Cases and controls undergo MRI including T1, T2 and MP2RAGE structural imaging, task (Go/NoGo) and resting-state MRI, and diffusion-weighted imaging (DWI). Concentrations of PM are being assessed using land use regression models, which incorporate data from air monitoring networks, dispersion models, and characteristics of roads and other land cover types. The estimated concentrations will be assigned to prenatal and postnatal residential and preschool/school addresses of all study subjects. We will assess whether long-term exposure to outdoor PM affects brain function, structure, and connectivity in healthy children and those diagnosed with ADHD.

**Results and Discussion:** This comprehensive study will provide novel, in-depth understanding of the neurodevelopmental effects of air pollution. f

## Introduction

### Background

Air pollution has received increased global public attention in recent years as presenting a major risk to human health (Manisalidis et al. 2020). According to estimates of the World Health Organization (WHO), ambient air pollution accounts for 4.3 million deaths per year (World Health Organization 2016). Exposure to outdoor particulate matter (PM) with aerodynamic diameter <2.5 µm (PM_2.5_) is the fifth-ranking risk factor for mortality worldwide (Cohen et al. 2017). Epidemiological, pathophysiological, and animal model studies provide extensive evidence that short- and long-term exposure to particulate air pollution causes cardiovascular diseases (Al-Kindi et al. 2020; Rajagopalan et al. 2018), contributes to respiratory health deterioration and allergy development (Bowatte et al. 2015; Lu et al. 2020a), and leads to lung cancer (Yin et al. 2017; Yu et al. 2018). Organic compounds and trace metals in PM may harm human health throughout the life course, beginning in utero (Achilleos et al. 2017; Bove et al. 2019; Lu et al. 2020b; Myhre et al. 2018).

PM has been found incorporated within human brain tissue. Most likely, it enters via the olfactory and the gastrointestinal nerves, and is associated with abnormal protein aggregation in the brainstem (Calderon-Garciduenas et al. 2020; Maher et al. 2016). In studies investigating the effects of PM using rodent models, inflammation and histopathological changes in the brain (Fu et al. 2020), and behavioural alterations (Cserbik et al. 2020; Nephew et al. 2020) have been attributed to PM exposure. Two recent studies on mice and rats showed that astrocyte function and mitochondrial activity in the cortex was severely affected by PM; larger effects were observed for exposure to smaller particle sizes (Araujo et al. 2019; Di Domenico et al. 2020). Some studies have shown that airborne PM can affect the structure and the functioning of the human brain (Herting et al. 2019), but the result of this research is far from conclusive. Also, it remains unclear whether exposure to PM can increase inattention, hyperactivity (Lu et al. 2020b), and impulsivity symptoms (Guxens et al. 2018), or impact on attention deficit hyperactivity disorder (ADHD) (Aghaei et al. 2019).

Emissions of many air pollutants in the European Union (EU) have decreased over the past three decades, and air quality is gradually improving. In particular, between the years 2000 and 2014, significantly decreasing trends in annual average concentrations of PM_10_ and nitrogen dioxide (NO_2_) were reported (Annesi-Maesano 2017; Guerreiro Cristina BB et al. 2014). Despite these improvements, EU urban populations are exposed to PM levels that exceed the WHO limits for the protection of human health (Sicard et al. 2021). Between 2000 and 2010, the daily limit value for PM_2.5_ and PM_10_ concentrations in the EU were exceeded 16 to 52% and 18 to 44%, respectively (Sicard et al. 2021). These excess PM levels are mostly observed in the eastern EU countries of Bulgaria, Czech Republic, Slovakia, and Poland (Guerreiro Cristina BB et al. 2014; Sicard et al. 2021).

Most of the PM pollution in Poland results from combustion of coal and other fossil fuels in power and heat generation, and from traffic emissions (Khomenko et al. 2021; Traczyk and Gruszecka-Kosowska 2020). Estimates of the mortality attributable to PM_2.5_ based on air quality monitoring data for Poland, have demonstrated that the urban Polish population experiences an unduly high impact of particulate air pollution on their health (Badyda et al. 2017). Annual deaths attributable to air pollution in Poland were estimated at 39,800 for the year 2000, increasing to 47,300 deaths for the year 2017 (Holnicki et al. 2017).

The overall objective of the NeuroSmog study is to assess whether long-term exposure to outdoor PM affects brain function, structure, and connectivity in both healthy children and those diagnosed with ADHD. Primarily, we will focus on the effect of PM on neural systems for attention and inhibitory control. In addition to neuroimaging and general intelligence quotient (IQ), impacts of PM on these systems will be investigated using behavioural tests. To meet the study aim, we assembled a multidisciplinary team of experts comprising air pollution modellers, environmental epidemiologists, neuroscientists, and clinical psychologists.

## Methods

### Study area

We selected towns in Poland based on their levels of particulate air pollution and population size that are located within two hours’ drive to the MRI scanning centre in Kraków. The particulate air pollution level of each town was classified as high, medium, or low based on the population-weighted median air pollution level inferred from 1km x 1km interpolated PM_2.5_ data for the year 2015 (European Environment Agency 2018). We chose small and large towns across different levels of air pollution to minimize urbanicity-related confounding. Towns with more than 90,000 inhabitants were classified as large; others were classified as small. Accordingly, we selected 18 towns (Table 1, Figure 1).

**Table 1.**
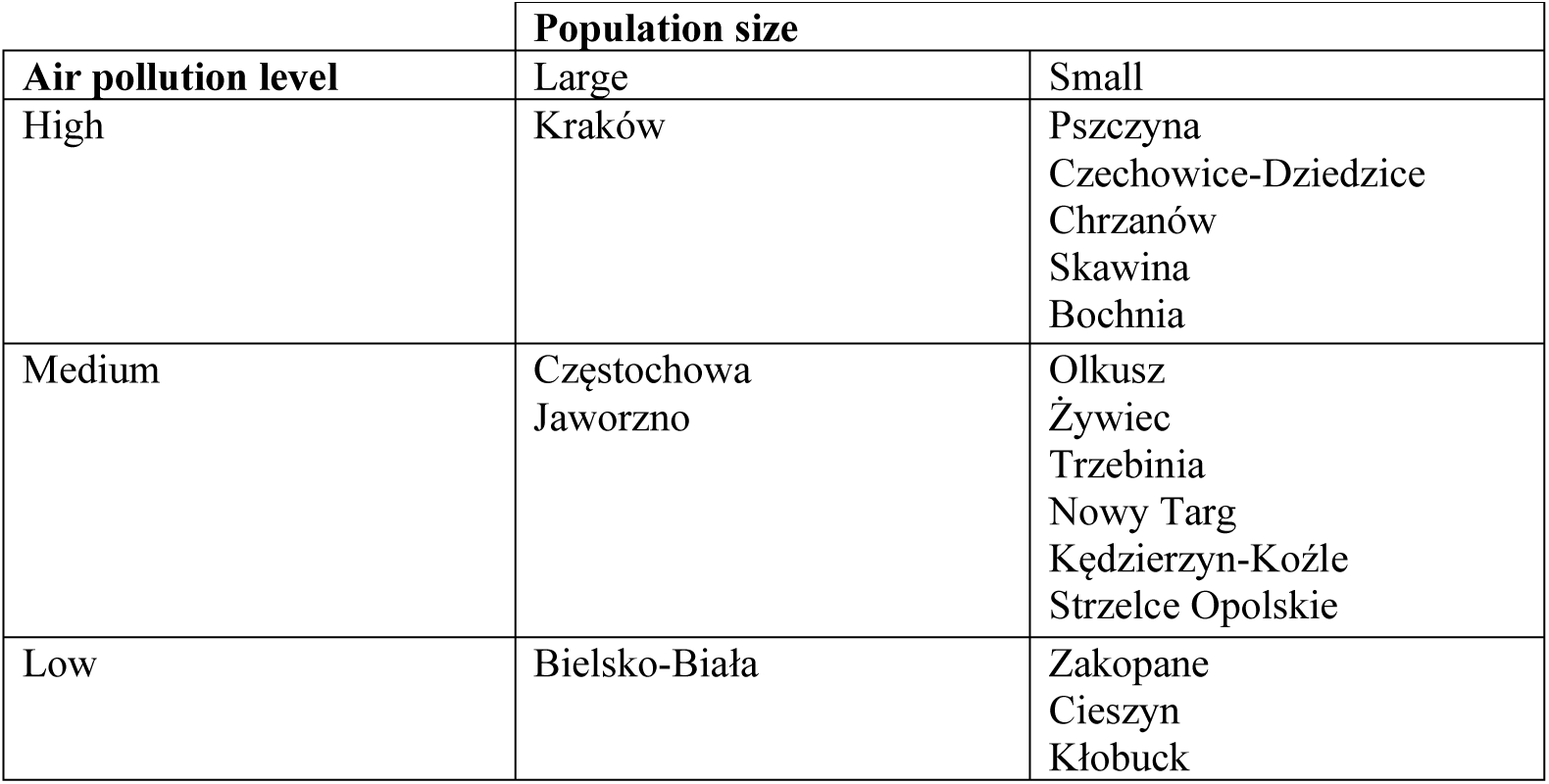
NeuroSmog study towns by type

| Air pollution level | Population size |  |
| --- | --- | --- |
|  | Large | Small |
| High | Kraków | Pszczyna<br>Czechowice-Dziedzice<br>Chrzanów<br>Skawina<br>Bochnia |
| Medium | Częstochowa<br>Jaworzno | Olkusz<br>Żywiec<br>Trzebinia<br>Nowy Targ<br>Kędzierzyn-Koźle<br>Strzelce Opolskie |
| Low | Bielsko-Biała | Zakopane<br>Cieszyn<br>Kłobuck |

**Figure 1.**
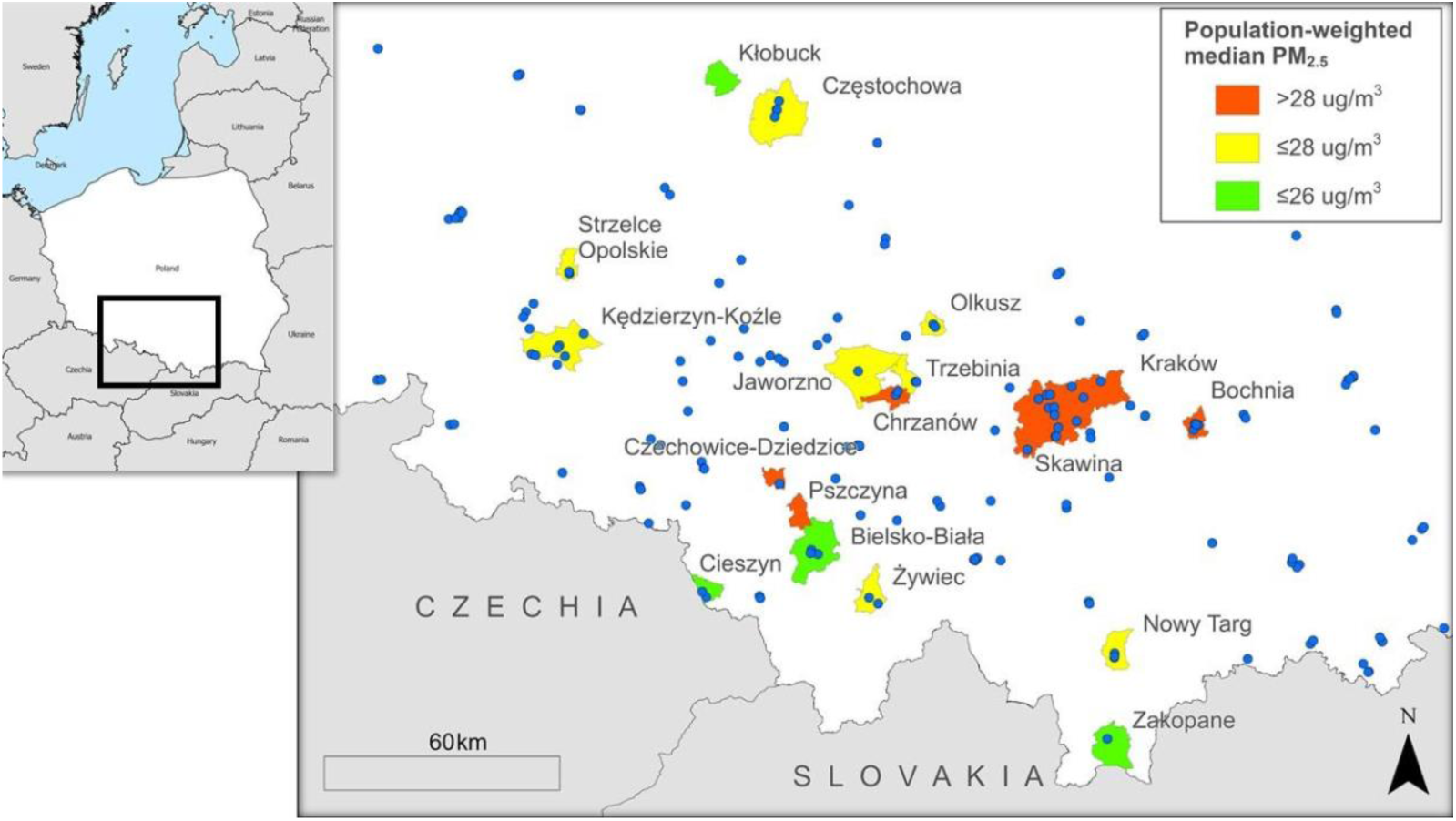
NeuroSmog study towns coloured according to their population-weighted median PM_2.5_ (2015) levels, as well as the locations of the air monitoring stations used for air pollution modelling (blue dots), and location of Poland on the map of Europe (study area highlighted by a black square).

We are recruiting comparable numbers of children (cases and controls) for each combination of air pollution level and town population size. Within each such combination, the number of children recruited per town is approximately proportional to the town’s population size (Table 1).

### Study design and study population

Recruitment of study participants began in October 2020, with the aim of 800 children completing all study procedures by the end of 2022. We are recruiting one ADHD case (intended number N=267) per two population controls (intended number N=533).

Cases with ADHD diagnosis according to the 11^th^ revision of the International Classification of Diseases and Related Health Problems (ICD-11) (World Health Organization 2019) are recruited from clinics in the study towns.

We use a two-step model to verify and eliminate ADHD diagnosis in cases and controls, respectively (see psychological evaluation section). In the first step, a comprehensive psychological assessment is administered and evaluated by local psychologists. In second step, three consultant clinical psychologist review the assessments to verify the diagnosis according to the ICD-11. Cases that do not meet the ICD-11 criteria are excluded from the study; controls who meet the ADHD diagnostic criteria are recruited into the case arm.

All study participants are native Polish speakers, aged between 10 and 13 years, have average or above average intelligence (i.e. attend non-specialised schools), are in grades IV - VI, and attend school in the selected towns. Exclusion criteria for all are diagnosed intellectual disability, neurological or comorbid psychiatric disorders, other serious medical conditions, as well as contraindications to MRI. Children born before the 35^th^ week or after the 40^th^ week of gestation, or with birth weight <2500 g or with an Apgar score <8 are also excluded. As we are interested in long-term effects of air pollution, children that were previously residing outside of Poland for at least a year are also excluded, as are children whose legal guardians are not fluent in Polish. The NeuroSmog study was approved by the Ethical Committee of the Institute of Psychology, Jagiellonian University, Kraków, Poland (# KE_24042019A; Clinical Trials Identifier NCT04574414).

A team of local psychologists with at least five years of clinical experience was recruited to enlist potential cases and perform psychological evaluation of all children. Prior to beginning their work, all psychologists received accredited training in the use of psychological tests utilised in the study (see section “Psychological testing”). They were also trained in the administration of two behavioural tests (see section “Behavioural tasks”), as well as in data protection and privacy laws.

Population controls are recruited from randomly selected larger non-specialized primary schools within each of the study towns (one to five schools per town). Lists of candidate schools were retrieved from the Boards of Education (Kuratoria Oświaty). Within each of the selected schools, classes are randomly selected in grades IV - VI and within each class, one to five pupils are selected at random. The sampling is defined *a priori* using randomized lists representing a maximum number of classes and pupils within the same grade in each school and class, respectively. The randomization was implemented using the *sample*.*int()* function in R statistical software (R Core Team 2018).

Headteachers of selected schools are contacted and asked to support the study’s recruitment campaign and sign a formal collaborative agreement. Participating schools are promoted on the project website, are offered workshops on neurodevelopment and on the study results. Consent for contact from the legal guardians is obtained by the schools. Families who give their consent are contacted by telephone by research assistants. The assistants explain the study aims and procedures, confirm study eligibility, and invite the legal guardians to participate in the study.

If a school does not agree to participate, the next school in the randomized list of schools is contacted. If the legal guardians of a selected child do not consent for contact, the next family from the randomized list of pupils is contacted. Where insufficient numbers of pupils are recruited in each grade, additional pupils are selected from the next class in the randomized list of classes (for that grade) using the same procedure until the desired number of pupils are recruited.

### Study timeline overview and progress

All psychological and behavioural assessments are performed during three meetings with the psychologist. Each meeting lasts about two hours, and both the child and their legal guardian are present. At the final meeting, the results of the assessments are explained to the child and their guardian and include a formal psychological evaluation. The MRI scan is completed in Kraków within three months of the psychological evaluation. After the MRI session, families are rewarded with Sodexo vouchers equal 270 PLN (about 60 EUR at the time of writing) and are reimbursed for travel expenses. The children receive t-shirts printed with MRI images of their brains.

Figure 2 illustrates the progress in the recruitment of controls the end of first study year (June 2021).

**Figure 2.**
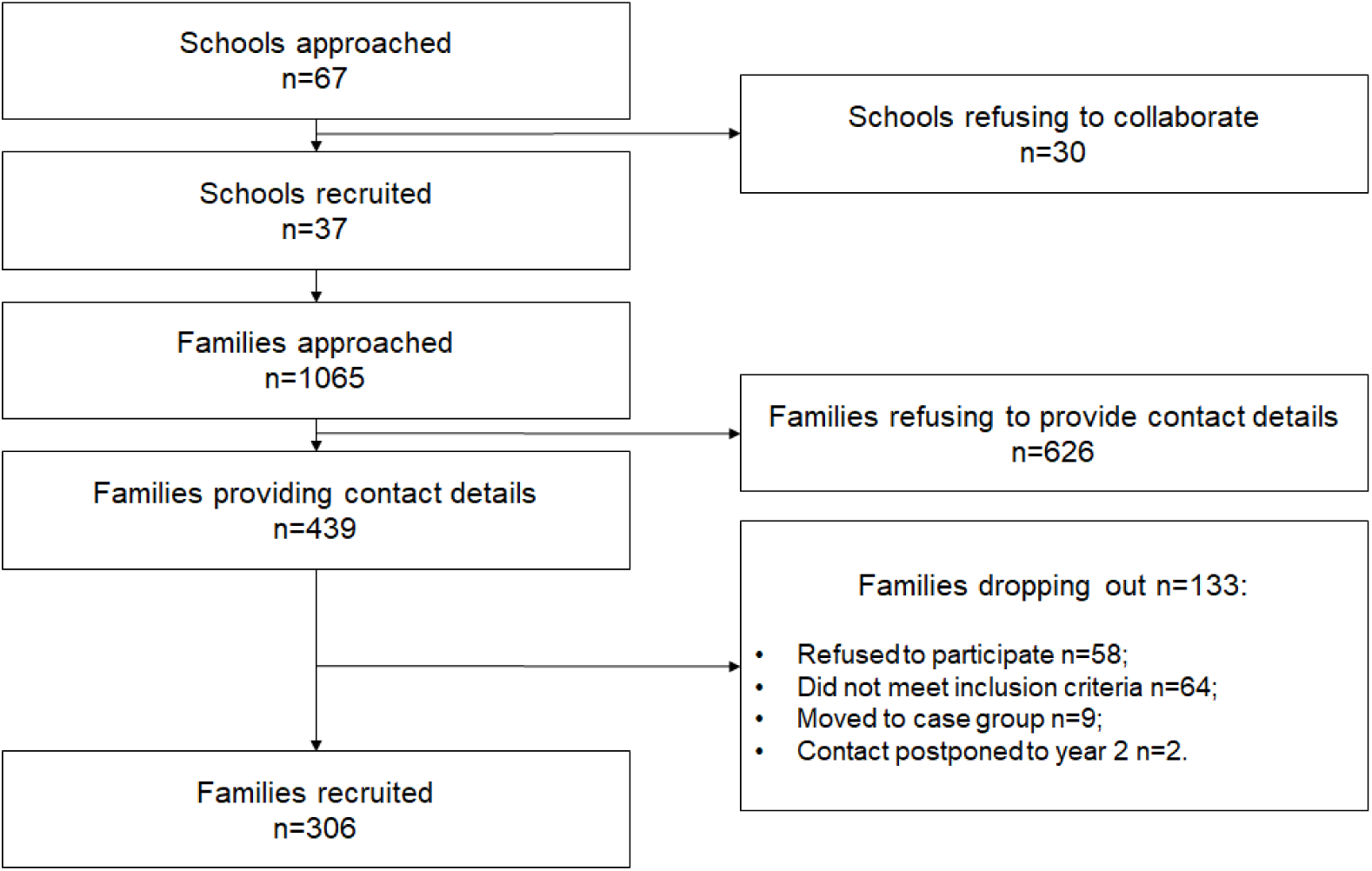
Flow chart describing the progress in the recruitment of controls at the end of the Year 1 (October 2020 to June 2021).

A total of 370 children had undergone psychological evaluation, of whom 236 had also completed the MRI scan. Figure 3 depicts the number of cases and controls tested in each town by end of August 2021.

**Figure 3.**
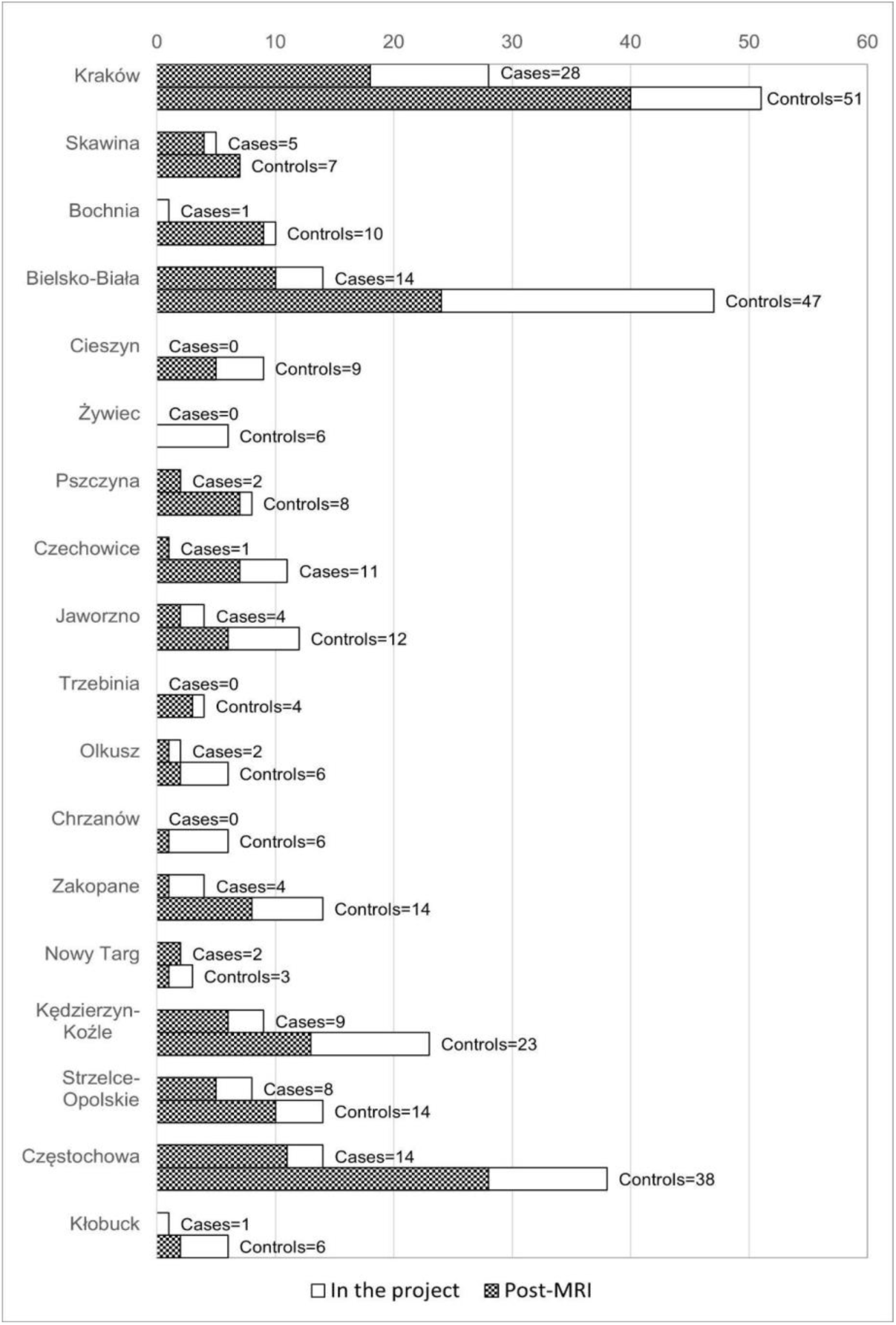
Town-level control-case specific progress of testing children in the project as of 09.09.2021.

**Figure 4.**
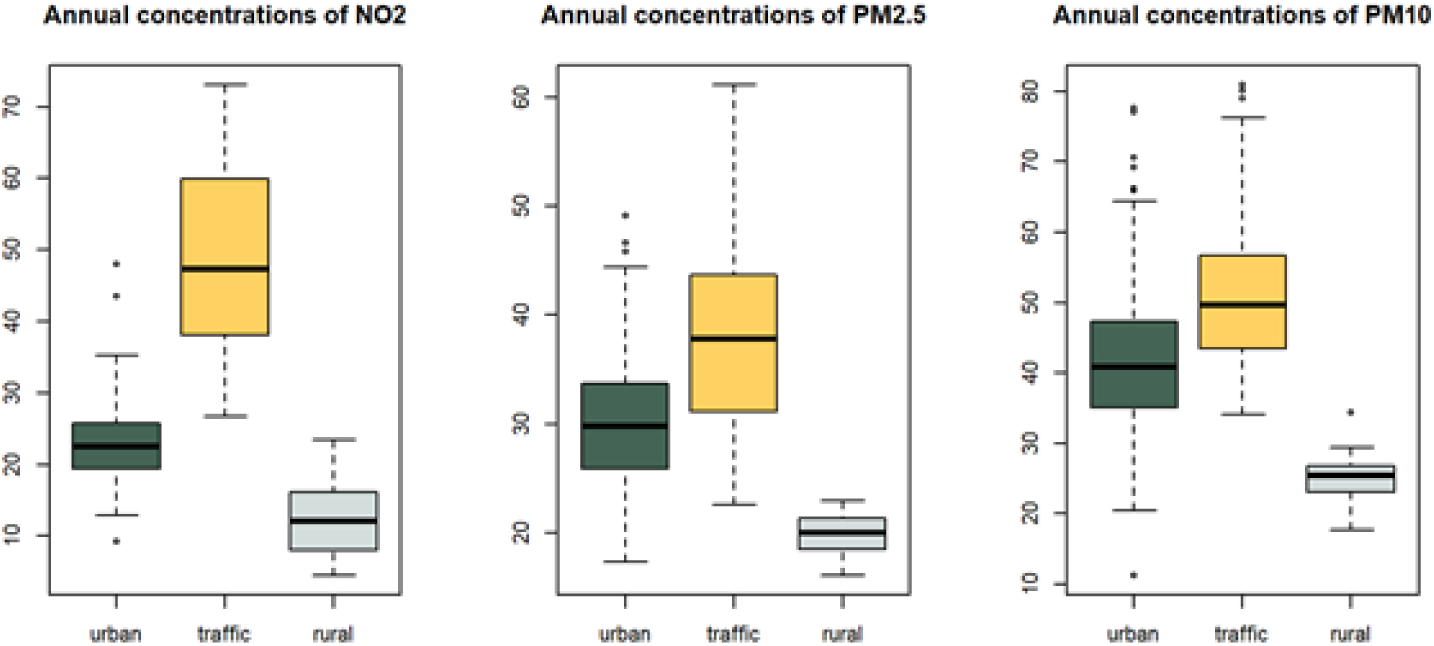
Boxplots illustrating annual means of daily measured PM_2.5_, PM_10_, and NO_2_ air pollutants grouped by type of station (urban, traffic, rural) from 2007 to 2019, based on monitoring stations in the study area. Between 2007 to 2019, the annual mean values of air pollutants concentrations at the monitoring stations ranged from 4.5 to 73.1 µg/m^3^ for NO_2_, from 11.2 to 80.9 µg/m^3^ for PM_10_ and from 16.1 to 61.1 µg/m^3^ for PM_2.5_.

### Data management

Behavioural testing is run on HP laptops with 15.6” screens, set up identically for each psychologist. The laptops are encrypted using BitLocker and synchronised with a secure OneDrive cloud folder at the Jagiellonian University that collects raw data from every test performed.

To minimise the impact of the study on the environment and to reduce transcription from paper to computer, the psychological data are - wherever possible - collected digitally using online questionnaires implemented in survey tools developed by Qualtrics (https://www.qualtrics.com).

The raw data are processed and prepared for import in R and Python. To allow easy data selection and extraction of variables of interest by data analysts, a secure user website will be implemented that will interface with an SQLite database.

### Study-specific questionnaires

Two questionnaires have been developed for the study: a General Questionnaire, and an Address Questionnaire. The General Questionnaire is a Qualtrics-based tool filled in by a legal guardian that collects data on confounders, effect modifiers, and additional exposures. Some items were adopted from the existing studies (Dzhambov et al. 2018a; Dzhambov et al. 2018b; Heinrich et al. 2017; Kuiper et al. 2020), and some of the items were invented for the purpose of NeuroSmog. Briefly, information on socio-demographics, pregnancy and early life, general health, habits, academic performance, home, and neighbourhood environments of the child is being collected (see Supplement A for a paper-adopted version of the questionnaire). Non-response options are always presented as a variation of “not applicable”, “don’t know”, “don’t wish to respond”, and at the same time, all responses are forced. General Questionnaire was validated on a small sample of volunteers.

The Address Questionnaire collects full pre-natal and post-natal residential history of the study participants, as well as preschool and school addresses, hours spent in preschools and at schools, and transportation mode used (see Supplement B). It is a pencil-and-paper questionnaire. The collected addresses are being geocoded and will be used to assign air pollution estimates, as well as other geographical exposures.

### Psychological evaluation

Cognitive functioning is examined using the Standford-Binet Intelligence Scales, 5^th^ edition (SB5; (Roid et al. 2017) and tests from the Diagnostic Battery for Cognitive Functions Evaluation (PU1; (Borkowska et al. 2015). These tests measure attention, working memory, and executive function, which represent domains relevant to both, classroom learning and ADHD (Holmes et al. 2020). Guardians complete the polish adaptions of the Third Edition of Conners’ Rating Scales (Wrocławska-Warchał E and Wujcik R 2018) and Child Behaviour Checklist (CBCL) for Ages 6-18 (Achenbach TM and Rescorla LA 2007), providing information on ADHD and other externalising and internalising behavioural issues. Children complete the polish adaption of Youth Self-Report (YSR) (Wolańczyk T 2002), which is a child-completed version of CBCL (Achenbach TM 2009; Achenbach TM and Rescorla LA 2007).

Social functioning is assessed using two questionnaires. The first is the Polish adaptation of the FACES-IV questionnaire (SOR) (Margasiński 2015), which measures family function, communication and family satisfaction. The SOR is completed by both guardians and children. The second is the Polish Siblings Relationship Questionnaire (KRR) (Lewandowska-Walter et al. 2016), which is completed only by children with siblings.

### Behavioural tasks

The attention network task (ANT) measures the efficiency of all three attentional networks - orienting, alerting, and executive functioning (Rueda et al., 2004). ANT demonstrates high immediate test-retest reliability, and scores are not correlated across individuals, suggesting that each network efficiency can be measured somewhat independently (Fan et al. 2002). We use the children’s version of ANT with timing and task stimuli operationalised identically to those used in Rueda et al. (2004) with the exception that, since our subjects are older, cue duration is set to 100 ms (as in the adult version).

The Continuous Performance Test (CPT) is a variant of the Go/NoGo task which allows quantification of deficits in attentiveness, impulsivity, activation/arousal, and vigilance. The CPT has high three months test-rested reliability (Conners CK et al. 2018) and has been validated in children with ADHD and in healthy controls (Seidel and Joschko 1990). During the task, geometric figures appear (250 ms) on the computer screen in 18 consecutive blocks of 20 trials (360 trials total). The geometric figures are identical to those used subsequently in the functional MRI (fMRI) variant of the Go/NoGo task. The inter-stimulus interval (ISI) is set to either 1, 2 or 4 seconds during each block. Participants are asked to press the spacebar in response to each letter presentation, except for the square and the circle (Conners et al. 2003). The proportions of Go to NoGo trials are set at 80% and 20%, respectively (Shaked et al. 2020).

All children complete practice runs of both tests until they performance reaches 90% accuracy.

### Neuroimaging

All neuroimaging data are acquired at the Małopolska Centre of Biotechnology, Jagiellonian University in Kraków, Poland, on a Siemens MAGNETOM Skyra 3T MRI scanner using a 64-channel head coil. Participants are familiarised with the scanner and trained to remain still using a mock scanner (pstnet.com). During the scan, head movements are constrained by inflatable Pearltec pillows (www.pearl-technology.ch).

The protocol includes T1-weighted (T1-w), T2-weighted (T2-w) and magnetization-prepared 2 rapid acquisition gradient echo (MP2RAGE) structural images, diffusion-weighted imaging (DWI), two rsMRI and two task fMRI runs (see Table 2 for detailed acquisition parameters and order). The scanning protocol, including instructions, lasts for one hour. The acquisition sequences for T1-w, T2-w and fMRI neuroimaging data were adopted from the Adolescent Brain Child Development project (Casey et al. 2018).

**Table 2.** Neuroimaging scanning parameters

| Sequence | Matrix | Slices | FOV | % FOV phase | Resolution (mm) | TR (ms) | TE (ms) | TI (ms) | Flip angle (deg) | Parallel Imaging | Multi band acceleration | Phase partial fourier | Diffusion directions | b-values | Acquisition time |
| --- | --- | --- | --- | --- | --- | --- | --- | --- | --- | --- | --- | --- | --- | --- | --- |
| **T1-w** | 256×256 | 176 | 256×256 | 100% | 1.0×1.0×1.0 | 2500 | 2.88 | 1060 | 8 | 2× | Off | Off | N/A | N/A | 06:09 |
| **T2-w** | 256×256 | 176 | 256×256 | 100% | 1.0×1.0×1.0 | 3200 | 565 | N/A | Variable | 2× | Off | Off | N/A | N/A | 05:34 |
| **fMRI** | 90×90 | 60 | 216×216 | 100% | 2.4×2.4×2.4 | 800 | 30 | N/A | 52 | Off | 6 | Off | N/A | N/A | 2*4.11 (task fMRI)<br>2*6.08 (rsfMRI) |
| **Diffusion** | 104×104 | 72 | 210×210 | 100% | 2.0×2.0×2.0 | 3800 | 101 | N/A | 78 | Off | 3 | 6/8 | 117 | 0 (10 dirs)<br>500 (18-dirs)<br>1250 (36-dirs)<br>2500 (53-dirs) | 07:31 |
| **MP2RAGE** | 256×256 | 176 | 256×256 | 100% | 1.0×1.0×1.0 | 5000 | 3 | 700 | 4 | 3x | Off | Off | N/A | N/A | 08:22 |

The DWI acquisition sequence was adapted from the UK Biobank (www.ukbiobank.ac.uk) to fit the study scanner and includes multiple b-value volume acquisitions. For the fMRI and DWI acquisitions we use multi-band accelerated EPI pulse sequences developed by the Center for Magnetic Resonance Research at the University of Minnesota (Feinberg et al. 2010; Xu et al. 2013). Magnetic susceptibility distortions will be corrected using collected references volumes with an inverted phase encoding direction.

The T1-w sequence provides a better contrast-to-noise ratio in white matter and is used for cortical and subcortical segmentation of the brain. The T2-w sequence allows the discrimination of structural differences in cerebral fluid-filled regions. Combined use of T1-w/T2-w allows the generation of ratio-based cortical myelin maps (Glasser and Van Essen 2011). It also provides the anatomical reference for rsMRI and task fMRI data, including anterior-posterior commissure alignment. For improved quality, the T1-w and T2-w protocols include volumetric navigators for prospective motion correction (Tisdall et al. 2012). MP2RAGE achieves a spatially uniform contrast (Marques et al. 2010) and is used for morphometric analyses of brain anatomy and for construction of cortical myelin maps. DWI enables visualisation and characterisation of white matter tracts (Assaf and Pasternak 2008). rsMRI imaging, allows to infer the intrinsic organization of large-scale brain networks (Buckner et al. 2013).

Throughout the structural, MP2RAGE and DWI acquisitions, participants watch neutral in valence movies (e.g., nature documentaries on the life of birds); throughout the rsfMRI acquisition, participants are instructed to stay awake, to keep still, and to blink normally while looking at a fixation cross.

Finally, task fMRI analyses can target task-dependent whole brain activity and functional connectivity (Friston 2009; Logothetis 2008). Based on our hypotheses, we chose an fMRI task that measures response inhibition as a measure of executive functioning, as inhibitory control is positively associated with cognitive flexibility and problem-solving skills and thus represents an important functional domain during brain maturation (Dowsett and Livesey 2000). In addition, diminished inhibitory control is associated with disorders of impulse control, including ADHD (Eme 2007; Romer et al. 2009). This task is identical to the conditioned approach response inhibition (CARIT) task used in the Human Connectome Project - Development (HCP-D) (Somerville et al. 2018) and is a variant of the Go/NoGo task. The task allows mapping of differential neuronal activity when response inhibition demands are high (NoGo trials) as compared to free prepotent motor execution (Go trials) (Conners CK et al. 2018). The task uses a rapid event-related fMRI design with jittered inter-trial intervals (1000-4500 mm) and randomized inter-target intervals to optimize statistical efficiency (Dale 1999). During each run, participants view shape stimuli (n=92) and are instructed to press a button as quickly as possible to every shape (Go; n=68) except for a circle and square (NoGo; n=24) (Newman et al. 2016; Somerville et al. 2018).

### Air pollution exposure assessment

Three health-relevant air pollutants are considered for this study: PM_2.5_ (Particulate Matter < 2.5 μg/m^3^), PM_10_ (Particulate Matter < 10 μg/m^3^) and NO_2_ (Nitrogen Dioxide). Air pollution measurements are collected in the air quality monitoring stations by the Polish Chief Inspectorate of Environmental Protection (Air Quality Portal 2021). The locations of the monitoring sites (n=179) in the study area are illustrated in Figure 1. Annual and monthly grids of air pollutant concentrations will be created for the study area for the period 2007 to 2022 using hybrid land use regression (LUR) models, similar to de Hoogh and colleagues (de Hoogh et al. 2013; de Hoogh et al. 2018). Interestingly, the research evaluating the performance of various regression models, regularisation techniques and machine learning methods, for air pollution spatial modelling has shown relatively small differences in prediction of accuracy in air concentration between various algorithms (Chen et al. 2019).

The LUR models will be estimated and validated using measurements from the air quality stations for each of the air pollutants and for each year separately and will also include monthly LUR models for the prenatal period.

Predictor variables for the LUR models are summarized in Table 3. We will calculate atmospheric dispersion of air pollutants according to meteorological conditions using methodology described in (Werner M et al. 2018; Werner M et al. 2019), providing 1hour concentrations of pollutants. We will use satellite images to inform about the land use changes. Data processing will be completed with QGIS geographic information system (GIS) software (QGIS Development Team 2021).

**Table 3.** Predictor variables for the air quality LUR modelling

| Type | Source | Main indicators | Characteristics |
| --- | --- | --- | --- |
| Emissions | National Centre for Emissions Management (National Centre for Emissions Management 2021), regional data from the voivodeship authorities, emission sector maps | Traffic emissions of air pollutants<br>Residential emission of air pollutants | Daily data available, monthly/annual means incorporated in LUR |
| Land use | Corine Land Cover (Corine Land Cover, 2021) | Forest and wooded area<br>Residential area<br>Surface water | Annual data collection years: 2006, 2012, 2018 |
|  |  | Vegetation and agricultural area<br>Area under roads, rail and airport roads<br>Unused land, landfill, excavation<br>Remaining, undeveloped area |  |
| Road data | Database of Topographic Objects, 1:10 000 scale, nationwide (BDOT10k, 2021) | Type of road (e.g., highway, expressway, main road etc.)<br>Number of traffic lanes within one road | Data collection from years: 2019 / 2020 |
| Air quality | Atmospheric dispersion models (Werner M et al. 2019) | Estimates from the dispersion modelling | Hourly data available, monthly/yearly means incorporated in LUR |
| Meteorological conditions | Institute of Meteorology and Water Management National Research Institute | Temperature<br>Wind speed and direction<br>Precipitation<br>Relative humidity<br>Atmospheric pressure | Hourly data available, monthly/yearly means incorporated in LUR |

Land cover predictor variables are calculated for seven girds of the following resolution: ∼4km x 4km, 2km x 2km, 1km x 1km, 500m x 500m, 250m x 250m, 125m x 125m and 62.5m x 62.5m.

Exploratory and correlation analyses of average air pollutants concentrations from the monitoring stations and the predictor variables will be performed to assess temporal and spatial distribution patterns. Optimal resolution for each type of variable applied in the LUR air quality modelling will be determined based on physical interpretability and supported with statistical analyses, including machine learning. Finally, predictor variables will be included in LUR model only if they adhere to the predefined direction of effect.

Models will be validated with out-of-sample and cross-validations methods using data from the monitoring stations. Model assumptions will be checked through the analysis of residuals, as well as visual inspection of quantile-quantile plots. Log transformations will be tested to minimize the effect of non-normal errors and p-values will be calculated assuming normal distributions of errors. Kriging of air pollution concentrations will be performed as it is expected to significantly improve model performance of linear regression algorithms for spatial data (Chen et al. 2019). All LUR modelling will be conducted in R statistical software.

Finally, estimates resulting from the LUR models will be assigned to all residential and preschool/school addresses of each study participants. Prenatal, early postnatal, concurrent, and life-long air pollution estimates will be calculated.

## Conclusion

The primary objective of the Neurosmog study is to combine state-of-the-art multimodal imaging, psychological assessment, environmental epidemiology, and air pollution modelling to determine the impact of air pollutants on neurophysiological and behavioural outcomes in healthy children and in atypically developing children diagnosed with ADHD. NeuroSmog is based on an ethnically, racially, and culturally homogeneous Polish population, which reduces potential confounders.

NeuroSmog is the first large scale study to provide collateral measures of brain structure, function and connectome, with periodically stratified and lifespan (including prenatal) exposures to air pollutants, and comprehensive child psychological assessments. We use a single MRI scanner and a narrow age range of participants to eliminate multisite acquisition confounding and to reduce epiphenomena related to developmental and hormonal changes in children. Additionally, we have adapted our protocol to fit two other large neuroimaging studies, the ABCD and HCP-D, and we will make our data openly available to the scientific community providing a rich and easily-comparable resource. A particular novelty of NeuroSmog is the inclusion of an fMRI task that directly tests the functioning of neural systems for inhibitory control that are putatively affected by air pollution exposure.

The few neuroimaging air pollution studies conducted to date were done in cities with relatively clean air. Performing this work in southern Poland allows us to investigate the impact of air pollution at concentrations and ranges greater than studied previously, thus increasing the chances of discovering associations, if present. Compared to existing similar studies, the project is unique in terms of its case-control design. ADHD children are hypothesized to be more vulnerable to the effects of air pollution. Finally, high-resolution air pollution grids will be produced for the study area using state-of-the-art techniques. We are thus confident that the data generated by our project, when combined with appropriate analysis and interpretation, will yield new insights into factors that impact or alter neurodevelopmental trajectories in children.

## Challenges

The COVID-19 pandemic, and disruptions to work, movement and education that are associated with it, have greatly complicated the fieldwork component of the study. Even under normal conditions, the logistics of managing field work across multiple centres are challenging. In general, all fieldwork has required more effort than originally anticipated and is taking longer than planned. The pandemic has also impacted negatively on response rates: 50% of contacted schools refused to collaborate and outreach towards the remaining 50% remains challenging.

Use of MRI in research presents additional challenges. Some members of the public distrust MRI and on occasions it has been difficult to motivate families of both cases and controls to participate in the study. A mixed attitude towards research involving underaged participants, exacerbated by the recent rise of anti-vaccine sentiment, also poses a challenge.

Finally, as with all case-control studies, we anticipate that recall bias may affect our analyses, particularly in terms of early life factors and residences.

## Data Availability

All data produced in the present study are available upon reasonable request to the authors

## Abbreviations

ABCD: Adolescent Brain Child Development study
ADHD: Attention deficit hyperactivity disorder
ANT: Attention network test
CARIT: Conditioned approach response inhibition task
CBCL: Child Behaviour Checklist
CPT: Continuous performance test
DWI: Diffusion-weighted imaging
EU: European Union
FACES-IV: Family Adaptation and Cohesion Evaluation Scales
GIS: Geographic information system
HCP-D: Human Connectome Project – Development
IQ: Intelligence quotient
ISI: Inter-stimulus interval
LUR: Land use regression model
MP2RAGE: Magnetization-prepared 2 rapid acquisition gradient echo
MRI: Magnetic resonance imaging
NO_2_: Nitrogen dioxide
PM: Particulate matter
PM_2.5_: Particulate matter with aerodynamic diameter <2.5 µm
PM_10_: Particulate matter with aerodynamic diameter <10 µm
SOR: Skala Oceny Rodziny
WHO: World Health Organization
YSR: Youth Self-Report

## Acknowledgements

We would like to thank all children and their legal guardians for their participation in the study, partner schools for helping us to reach out to children assigned to the control group, psychologists for identifying children with ADHD and performing psychological testing on all children, and the study team for technical, logistic, administrative, and communication efforts. We would also like to thank the project’s Advisory Board members for the scientific support and advice. Finally, we would also like to thank Kees de Hoogh for assistance with air pollution modelling.

## Authors’ contributions

Iana Markevych designed the study, conceptualized the field work and is overseeing it, coordinating and executing the data management, consulting on the air pollution modelling, drafted the first version of the study protocol, revised the draft, and approved the final manuscript

Natasza Orlov is coordinating and executing the data management, drafted the first version of the study protocol, revised the draft, and approved the final manuscript

Munawar Hussain Soomro drafted the first version of the study protocol, revised the draft, and approved the final manuscript

James Grellier designed the study and obtained the necessary funds, is consulting on the air pollution modelling, revised the study protocol, and approved the final manuscript

Katarzyna Kaczmarek-Majer is responsible for the air pollution modelling, drafted the first version of the study protocol, revised the draft, and approved the final manuscript

Małgorzata Lipowska designed the study, conceptualized the field work and is overseeing it, revised of the study protocol, and approved the final manuscript

Katarzyna Sitnik-Warchulska designed the study, conceptualized the field work and is overseeing it, revised of the study protocol, and approved the final manuscript

Yarema Mysak conceptualized the field work and is overseeing it, is coordinating and executing the data management, drafted the first version of the study protocol, revised the draft, and approved the final manuscript

Clemens Baumbach designed the study, conceptualized the field work and is overseeing it, is coordinating and executing the data management, drafted the first version of the study protocol, revised the draft, and approved the final manuscript

Maja Wierzba-Łukaszyk conceptualized the field work and was overseeing it, was coordinating and executing the data management, revised of the study protocol, and approved the final manuscript

Mikołaj Compa conceptualized the field work and is overseeing it, is executing the data management, revised the study protocol, and approved the final manuscript

Bernadetta Izydorczyk designed the study, conceptualized the field work, and oversees it, revised of the study protocol, and approved the final manuscript

Krzysztof Skotak is responsible for the air pollution modelling, revised the study protocol, and approved the final manuscript

Anna Degórska is responsible for the air pollution modelling, revised the study protocol, and approved the final manuscript

Jakub Bratkowski is responsible for the air pollution modelling, revised the study protocol, and approved the final manuscript

Aleksandra Domagalik contributed to the design of the MRI acquisition protocol, revised the study protocol, and approved the final manuscript

Marcin Szwed is the principal investigator of the study and obtained the necessary funds, designed the study, conceptualized the field work and its oversight, drafted the first version of the study protocol, revised the draft, and approved the final manuscript

